# Faith- and Community-Led Community Posts in Zambia Close Gaps in Men by Expanding HIV Case Finding, Treatment and Viral Suppression Over 4 Years (April 2018 – March 2022) in the Context of COVID-19

**DOI:** 10.1101/2025.04.05.25325299

**Authors:** Gibstar Makangila, Francesca Merico, Susan Hillis

**Author notes:** **Corresponding Author:** Susan Hillis, PhD, MS. **E-mail addresses of authors:**.

## Abstract

**Introduction:** HIV testing and treatment coverage among men in Zambia remains suboptimal, partly due to lower healthcare-seeking behavior compared to women. To address this gap, Circle of Hope (CoH), a local faith-based organization funded by the President’s Emergency Plan for AIDS Relief (PEPFAR), developed the community post (CP) model. CPs are staffed by multi-disciplinary teams that provide HIV-testing, anti-retroviral therapy (ART), and viral load (VL) testing. This analysis assesses the impact of these CPs on HIV case finding, ART initiation, and VL suppression among men.

**Methods:** Routine program data reported to PEPFAR by CoH Zambia were analyzed for two periods: pre-COVID 19 (April 2018-March 2020) and during the pandemic (April 2020-March 2022). Indicators included the number of HIV tests conducted and positive results received (overall and for index testing specifically), ART initiation among men living with HIV (MLHIV), 12-month retention on ART, and the proportion of clients with VL suppression (<1000 copies/mL).

**Results:** Between April 2018 and March 2022, the number of CPs expanded from 9 to 38 across Zambia. Despite the challenges posed by the COVID-19 pandemic, men continued to represent a substantial share of clients: 49.5% to 41.4% of those tested, 38.5% to 42.0% of those diagnosed (p<0.001), and 38.9% to 43.3% of those initiated on ART (p<0.001). The number of men on ART rose from 3,320 (38%) to 13,580 (43%, p<0.000001). Retention and VL suppression among men also remained high, increasing slightly from 94% to 97% and 95% to 96%, respectively. Over 6,000 men were diagnosed through index testing services, with percent positivity consistently high, ranging from 54.4% to 61.1% (p<0.001).

**Conclusions:** The CoH model was successful at identifying and linking MLHIV to HIV treatment services and assisting them to achieve and maintain viral suppression even during the COVID-19 pandemic. Expanding this community-based model may help close HIV testing and treatment gaps among men and advance HIV epidemic control in Zambia and beyond.

## Introduction

Sub-Saharan Africa bears a disproportionate share of the global HIV burden, with over two-thirds of all people living with HIV – approximately 25.6 million – residing in the region [1]. To curb the epidemic, the Joint United Nations Programme on HIV/AIDS (UNAIDS) established the 95-95-95 targets for 2030: 95% of PLHIV should know their status, 95% of those diagnosed should receive antiretroviral therapy (ART), and 95% of those on ART should achieve viral load suppression (VLS) [2]. However, significant gaps persist, particularly among men (72% ART coverage), youth (52%), and children (57%) as of 2022 [3].

These disparities are interlinked, with recent evidence demonstrating that suppressing viral load in men can halve HIV incidence in women [4-6]. Men living with HIV (MLHIV) who remain unsuppressed face higher mortality risks [7-9], increasing the likelihood of orphanhood among their children.

Orphaned children are two to three times more likely to acquire HIV than their non-orphaned peers [10]. In Zambia, these challenges mirror global trends: as of 2022, the national HIV prevalence stood at 11.9%, with lower VLS among men (88% compared to 92% in women) and youth (80%), and a pediatric treatment gap of 30% [11-12].

Circle of Hope (CoH) Zambia, a local faith-based organization, identified key barriers to HIV service uptake: long wait and travel times, poor customer care, financial burdens (transport costs and lost wages); disjointed service delivery due to competition between providers and stakeholders, and HIV-related stigma [13-15]. These factors disproportionately affect men, who are less likely than women to engage with healthcare services.

To address these barriers, CoH Zambia, in collaboration with the U.S. Centers for Disease Control and Prevention (CDC), developed and implemented a faith- and community-led Community Post (CP) model. This model engages trusted faith leaders and community members to support the provision of HIV services at decentralized, unbranded, one-roomed community posts situated in convenient locations such as markets, bus stops, churches, mosques, and/or fishing camps. By engaging trusted faith leaders and community members, the CP model leverages Zambia’s religious landscape, where ~90% of people consider faith central to their lives [16-17], to enhance access, uptake, and continuation of essential HIV services.

This analysis evaluates the effectiveness of the CP model in improving HIV outcomes among MLHIV, including case identification, ART initiation, treatment retention, and viral suppression. Results are disaggregated by sex and analyzed across pre-pandemic and pandemic periods. Cost data for CP establishment are also presented to inform potential scale-up and sustainability.

## Methods

### The Community Post (CP) Model for Providing HIV Services

The CP model is a faith-and community-led approach designed to identify PLHIV, particularly men, and link them to HIV treatment services. The model directly addresses the barriers that men in accessing HIV services from conventional health facilities including time and resources constraints [18-22].

The CP model comprises three core components. The first component focuses on decentralized delivery of HIV services through CPs. Site selection is done in collaboration with local stakeholders to ensure that CPs are conveniently located within the communities where men live and work. Each CP is staffed by a multidisciplinary team that includes doctors, nurses, outreach workers, data clerks, and faith and community champions – trusted community members trained in client-centered care and compassionate service delivery. These champions leverage their deep knowledge of local social networks to identify individuals and families in need of HIV services and link them to the CPs for care. Outreach workers, trained by the Zambian government, deliver a wide range of HIV and primary healthcare services both at the CPs and within the community. Additional staff employed by the CPs include drivers, laboratorians who process diagnostic samples, pharmacists who dispense antiretrovirals and medications for opportunistic infections, and team leaders who provide ongoing supervision, training, and mentorship to ensure high-quality service delivery.

Each CP offers a standardized package of core HIV services including: HIV testing (with index testing for the sexual partner(s) and biological child(ren) of PLHIV); distribution of HIV self-test kits; same-day, community-delivered ART initiation and continuation (including multi-month dispensing and adherence counseling and support); viral load testing; and a range of HIV prevention services, such as pre- and post-exposure prophylaxis (PrEP/PEP), risk reduction counseling, condom distribution, and referrals for voluntary medical male circumcision. CPs also provide syndromic management of sexually transmitted infections (STIs); and cervical cancer screening and referrals. In addition, select CPs also offer broader primary healthcare services, either directly or through referral. These include screening for non-communicable diseases (e.g., hypertension, diabetes, obesity); support for mental health and substance abuse; maternal and child healthcare (e.g., antenatal services, immunizations, and teen pregnancy referrals); and violence prevention services for adolescents and young women.

The second component of the CP model centers on strategic partnerships with community and faith leaders, who play a pivotal role in the model’s success. These leaders help identify and select faith and community champions and outreach workers who are trusted, respected, and deeply familiar with the norms and networks in the local community. These leaders also support service delivery by publicly endorsing HIV services, which reduces HIV-related stigma and makes it easier for men to access these services [22-23]. The model’s collaborative approach—with strong links to government agencies, civic leaders, and the Ministry of Health—ensures alignment with national health priorities and strengthens the sustainability and scalability of the CP model [24].

The third component of the model emphasizes values-driven service delivery, grounded in the R.E.C.I.P.E. framework: Responsibility, Empathy, Compassion, Integrity, Passion, and Ethics. Each day begins with a team leader–led pep talk to reinforce these core values and motivate staff to deliver respectful, high-quality, client-centered care. Continuous supervision and mentoring—delivered both in person and via WhatsApp—help maintain fidelity to the model and ensure staff are providing a consistently high quality of care that encourages men to remain engaged in HIV services.

### Data Sources and Analysis

This study analyzed routine program data reported by CoH to the U.S. President’s Emergency Plan for AIDS Relief (PEPFAR) through the Monitoring, Evaluation, and Reporting (MER) framework.

Indicators included the number of HIV tests conducted and positive results received (overall and via index testing); the number of PLHIV initiated on ART; 12-month ART retention; and the proportion of ART clients with documented viral load suppression (<1000 copies/mL). Percent positivity (yield) was calculated as the proportion of HIV-positive results among total tests conducted.

Indicators were analyzed across two time periods: pre-pandemic (April 2018–March 2020) and during the first two years of the COVID-19 pandemic (April 2020–March 2022). Data were examined both overall and disaggregated by sex. Differences in proportions between time periods and between male and female clients were assessed using chi-squared tests, while disaggregated comparisons used Wilcoxon rank-sum tests. Statistical significance was defined as p<0.05, with p-values calculated under the assumption of asymptotic normality [25].

## Results

Between April 2018 and March 2022, the number of CPs expanded from 9 to 38 across four of Zambia’s ten provinces (Table 1). The vast majority (97%) were located in urban areas and staffed by a multi-disciplinary team comprising faith and community champions (68.4%), outreach workers (22.3%), clinical officers (5.4%), data clerks (3.3%), and nurses (0.6%).

**Table 1.** Expansion of Faith- and Community-Led Community Posts (CPs) Implemented by Circle of Hope, April 2018 – March 2022^1^.

|  | Apr 2018 – Mar 2019<br>(Year 1) | Apr 2019 – Mar 2020<br>(Year 2) | Apr 2020 – Mar 2021<br>(COVID) | Apr 2021 – Mar 2022<br>(COVID) |
| --- | --- | --- | --- | --- |
| Total number of CPs | 9 | 34 | 38 | 38 |
| Number of CPs by setting |  |  |  |  |
| -Urban | 9 | 33 | 37 | 37 |
| -Rural | 0 | 1 | 1 | 1 |
| CP staffing by cadre (n, %) |  |  |  |  |
| -Clinical officers (or medical doctors) | 9 (6.2%) | 34 (8.1%) | 38 (4.8%) | 38 (4.6%) |
| -Nurses | 3 (2.1%) | 3 (0.7%) | 4 (0.5) | 4 (0.5%) |
| -Faith and community champions<sup>2</sup> | 75 (51.7%) | 235 (56.2%) | 600 (73.3%) | 600 (72.6%) |
| -Outreach workers<sup>3</sup> | 52 (35.9%) | 136 (32.5%) | 152 (18.6%) | 152 (18.4%) |
| -Data clerks | 6 (4.1%) | 10 (2.4%) | 24 (2.9%) | 32 (3.9%) |

<sup>1</sup>Community posts were initially established by Catholic Relief Services with Circle of Hope as a sub-recipient. Beginning in September 2020, Circle of Hope took over the prime responsibility for establishing and overseeing these community posts.
<sup>2</sup>Faith and community champions are community members who are trained to provide value-based customer care using the RECIPE (Responsibility, Empathy, Compassion, Integrity, Passion, and Ethics) framework and to link individuals from their community in need of HIV services to CPs. In 2020, Circle of Hope received additional funds from PEPFAR to significantly expand the number of faith and community champions at each of the CPs based on the success of the model.
<sup>3</sup>Outreach workers are trained by the Zambia government to directly provide HIV services, including HIV rapid testing and adherence counseling, at the CPs.

The number of MLHIV receiving ART at CPs increased from 3,320 (38% of all PLHIV receiving ART at CPs in April—March 2019) to 13,580 (43% in April—March 2022; p<0.000001) (Figure 1). Despite the COVID-19 pandemic, the proportion of men receiving HIV services at CPs remained substantial.

**Figure 1.**
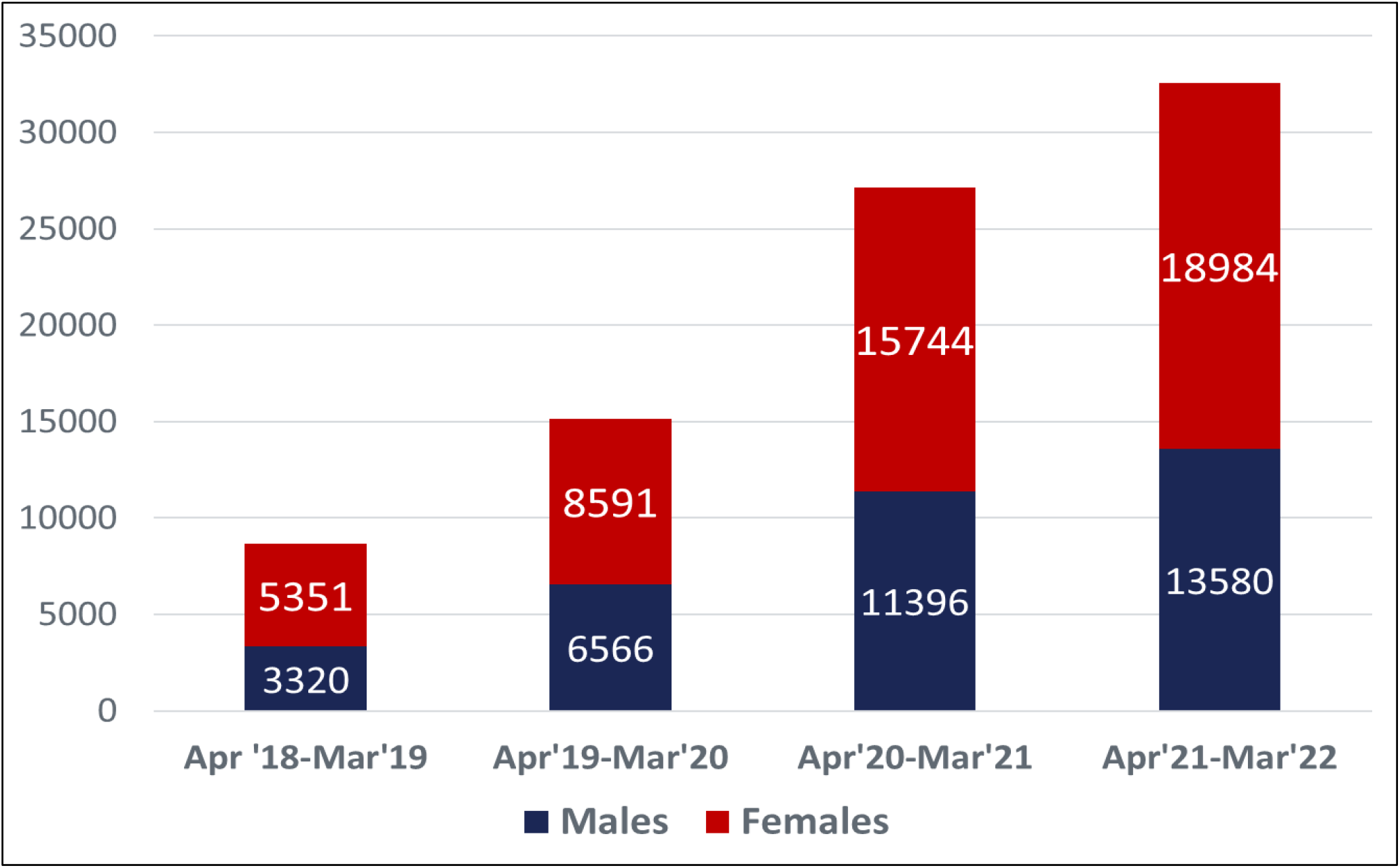
The Number of Men and Women Living with HIV Who Received Anti-Retroviral Treatment at the Faith- and Community-Led Community Posts Steadily Increased Before and During the COVID-Pandemic, April 2018 – March 2022, Zambia

Men accounted for 49.5% of clients tested pre-pandemic, compared to 41.4% during the pandemic. The proportion of HIV diagnosis among men rose from 38.5% to 42.0% (p<0.001), and ART initiation among MLHIV increased from 38.9% to 43.3% (p<0.001). Percent positivity also rose from 25% to 28% (p<0.001). Retention at 12 months and viral load suppression rates remained consistently high across all time periods, at or above 95%. Over 6,000 men were diagnosed through index testing services over the four-year period, with percent positivity ranging from 54.4% to 61.1% (p<0.001) (Tables 2 and 3).

**Table 2.**
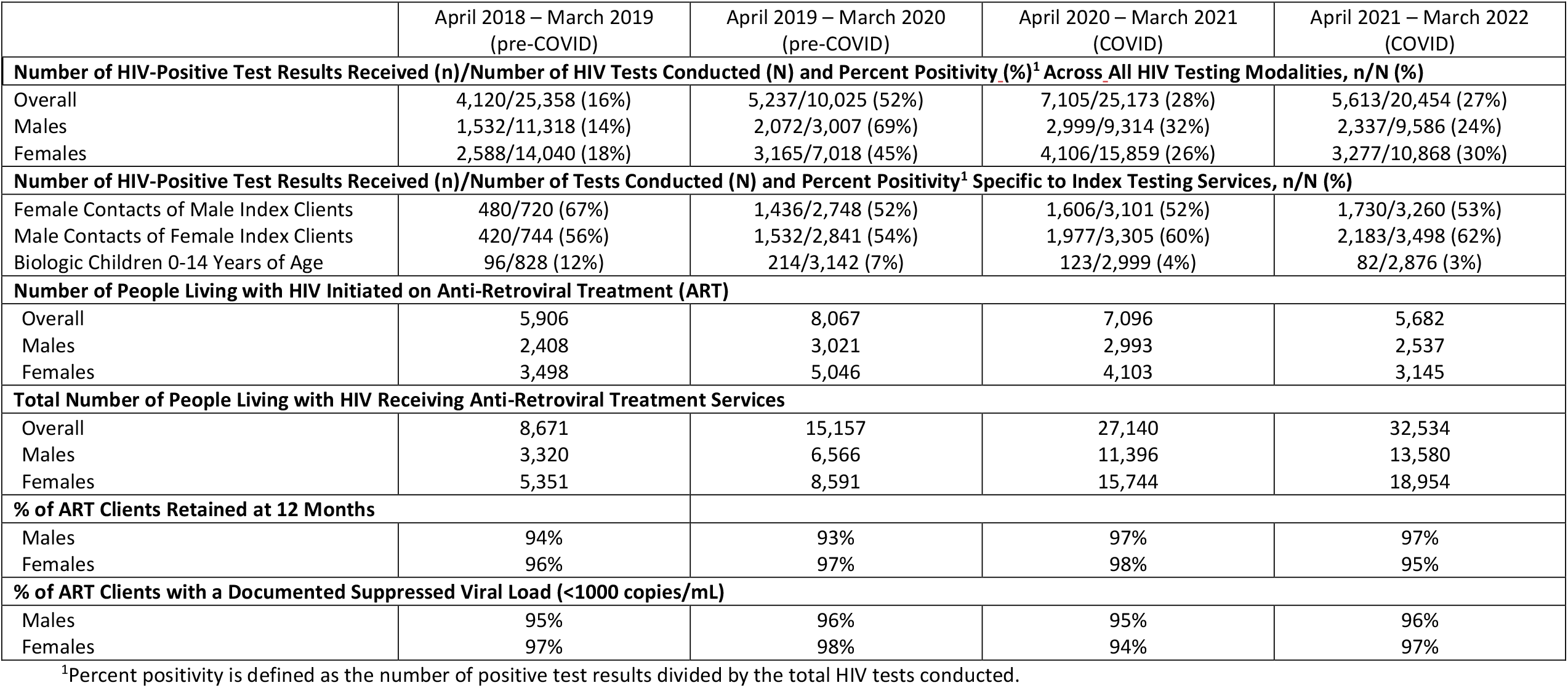
HIV Case Finding through Index Testing, Treatment Initiation, Retention at 12 Months, and Viral Load Suppression for Clients Served by Circle of Hope Faith- and Community-Led Community Posts, April 2018 – March 2022.

**Table 3:**
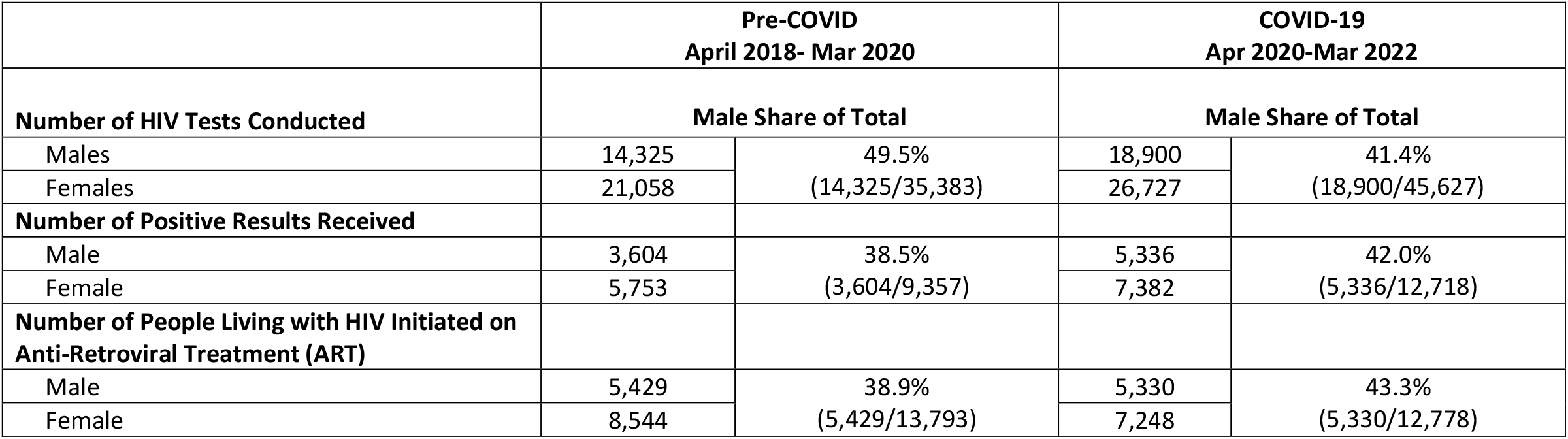
Male Share of the Total Numbers of HIV Tests Conducted, Numbers of Positive Test Results Received, and Numbers of People Living with HIV Initiated on Anti-Retroviral Treatment among All Clients Served by Circle of Hope Faith- and Community-Led Community Posts, April 2018 – March 2022.

To assess whether increases in treatment numbers were primarily due to client transfers from other facilities, CoH analyzed program data from April 2019 to March 2020 and April 2020 to March 2021. During 2019-2020, only 6.7% of the 8,067 clients who tested HIV-positive at CPs were transfers, while 93.3% were either newly diagnosed or returning to care after a treatment interruption. In 2020-2021, transfers accounted for 15.5% of the 7,096 HIV-positive clients, with the remaining 84.5% representing new diagnoses or re-engagement in care. These findings suggest that the observed increases in ART initiation were largely driven by new case identification and successful re-engagement of individuals previously lost to follow-up, rather than by transfers from other facilities.

In 2020, the Government of the Republic of Zambia began operating its own CPs. While these government posts were placed in the community, they lacked the other two components: strategic partnerships and value-based mentorship guided by the RECIPE framework. A comparative analysis from January to June 2021 showed that CoH-led CPs significantly outperformed government-supported CPs in reaching men with HIV services, achieving higher percent positivity (32.5% vs. 9.5%, p<0.001) and viral load suppression (94.8% vs. 90.4%, p<0.001) (Table 4). In response, CoH provided mentorship and support to government-run sites to implement all three components. Performance subsequently improved across key indicators including percent positivity (9.5% to 19.3%, p<0.0001) and viral load suppression (90.4% to 95.7%, p<0.01).

**Table 4.** Comparison of HIV Testing and Viral Load Suppression Outcomes Among Men Attending Circle of Hope-Sponsored Community Posts Compared to Government-Sponsored Community Posts, January–June 2021.

|  | Circle of Hope Posts | Zambian Government Posts<br>(before CoH mentoring)<sup>1</sup> | Zambian Government Posts<br>(after CoH mentoring) |
| --- | --- | --- | --- |
| **Number of Community Posts** | 38 | 67 | 67 |
| **Number Tests Conducted** | 4,095 | 2,992 | 13,321 |
| **Number of Positive Test Results** | 1,333 | 279 | 2,569 |
| **Percent Positivity<sup>2</sup>** | 32.5% | 9.5% | 19.3% |
| **Viral Load Suppression<sup>3</sup>** | 94.8% | 90.4% | 95.7% |

<sup>1</sup>Circle of Hope sites included all three strategies of the Community Post model: (1) health posts located within the community, (2) strategic partnerships with community and faith leaders, and (3) supportive supervision and mentoring using the R.E.C.I.P.E (Responsibility, Empathy, Compassion, Integrity, Passion, and Ethics) framework; government posts only included the community location before mentoring from CoH and all three components after mentoring.
<sup>2</sup>Percent positivity is defined as the total number of positive test results divided by the total number of HIV tests conducted.
<sup>3</sup>Viral load suppression is defined as <1,000 copies/milliliter.

Table 5 provides the operational expenses associated with operating a CP. Salaries accounted for most CP expenses (90%) followed by rent (3%), quarterly trainings (2%), utilities (2%), and supplies (2%). On average, each CP operated at an annual cost of approximately USD $20,000 per year, with staffing as the primary cost driver.

**Table 5.**
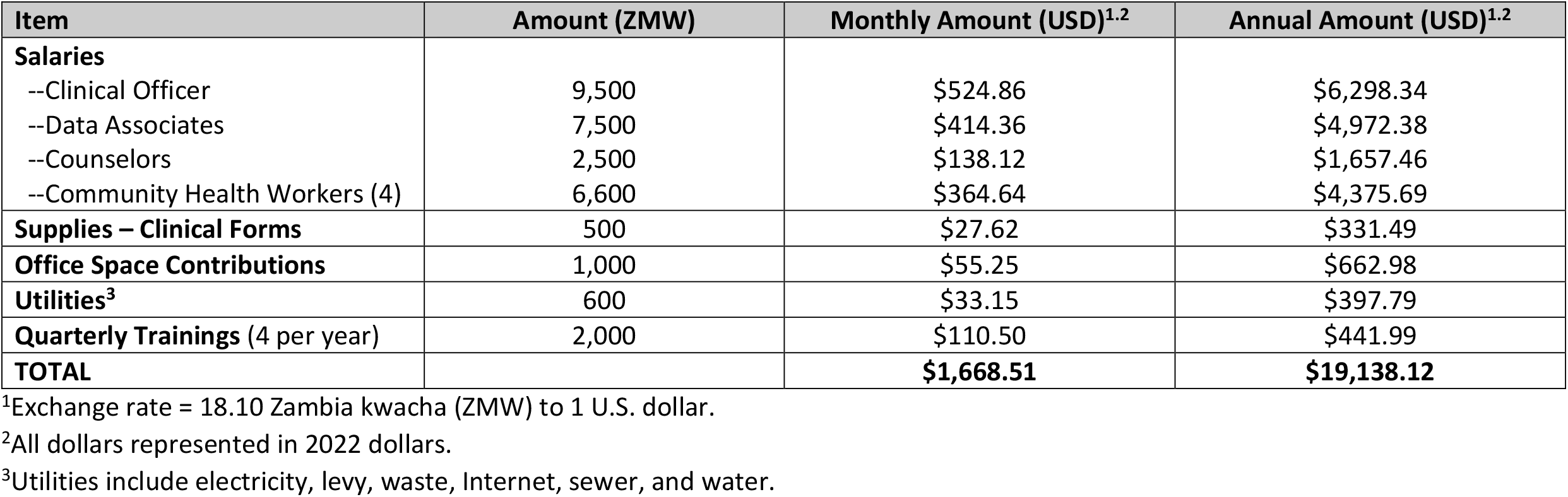
Operational Expenses for Faith- and Community-Led Community Posts, Circle of Hope, April 2018 – March 2022, Zambia.

| Item | Amount (ZMW) | Monthly Amount (USD)<sup>1,2</sup> | Annual Amount (USD)<sup>1,2</sup> |
| --- | --- | --- | --- |
| **Salaries** |  |  |  |
| --Clinical Officer | 9,500 | \$524.86 | \$6,298.34 |
| --Data Associates | 7,500 | \$414.36 | \$4,972.38 |
| --Counselors | 2,500 | \$138.12 | \$1,657.46 |
| --Community Health Workers (4) | 6,600 | \$364.64 | \$4,375.69 |
| **Supplies – Clinical Forms** | 500 | \$27.62 | \$331.49 |
| **Office Space Contributions** | 1,000 | \$55.25 | \$662.98 |
| **Utilities<sup>3</sup>** | 600 | \$33.15 | \$397.79 |
| **Quarterly Trainings (4 per year)** | 2,000 | \$110.50 | \$441.99 |
| **TOTAL** | | **\$1,668.51** | **\$19,138.12** |

<sup>1</sup>Exchange rate = 18.10 Zambia kwacha (ZMW) to 1 U.S. dollar.
<sup>2</sup>All dollars represented in 2022 dollars.
<sup>3</sup>Utilities include electricity, levy, waste, Internet, sewer, and water.

## Discussion

This study demonstrates that the CoH-led CP model significantly expanded HIV service access for men in Zambia—a group historically underserved in the HIV response. By leveraging trust, community presence, and flexible service delivery, the model effectively identified, initiated, and retained MLHIV on HIV treatment services, achieving high rates of viral suppression. Importantly, CPs not only sustained operations but expanded during the COVID-19 pandemic, maintaining strong outcomes across the HIV care cascade. Over four years, the number of men receiving ART at CPs quadrupled, from 3,320 to 13,579, highlighting the model’s scalability and resilience.

With index testing positivity rates ranging from 52% to 73% and viral load suppression rates between 95% and 96%, the effectiveness of the CP model compares favorably with other male-focused interventions. For example, the Lesotho Men’s Clinics – designed specifically for men with extended hours and non-stigmatizing, comprehensive services – reported HIV positivity rates 1.6% to 16% higher than national averages, along with a viral load suppression rate of 95.6% [26-27]. Similarly, a two-arm cluster randomized trial in African American churches in the United States found that men in intervention churches who received ongoing HIV messaging and onsite testing were nearly twice as likely to get tested (47%) compared to men who received only standard educational messages (28%) [28]. Together, these approaches underscore the value of tailored, community-based approaches that reduce HIV-related stigma and promote trust in closing gaps in the HIV treatment cascade for men.

Collectively, these findings highlight the power of community-tailored, stigma-reducing strategies—like the CP model—in improving HIV outcomes for men.

A key strength of the CP model is in its reimagining of HIV service delivery—from a traditional, facility-based approach to a decentralized, community-led model. In Zambia, men face unique barriers to accessing care such as poor customer service, income loss from time away from work, long wait and travel times, and HIV-related stigma [20]. The CP model directly addresses these challenges by engaging faith and community leaders to identify and train trusted local peers to deliver HIV services grounded in the RECIPE framework—a values-based approach emphasizing Respect, Empathy, Compassion, Integrity, Passion, and Ethics. Services at the CPs are intentionally streamlined—often completed in under 30 minutes—to minimize disruption to men’s daily responsibilities and encourage repeat engagement. By embedding trust, dignity, and convenience into the care experience, the CP model fosters both initial uptake and continued engagement in HIV services.

Comparative analysis showed that COH-supported CPs implementing all three core components of the model—decentralized service delivery in the community, community and faith engagement, and values-based mentoring through the RECIPE framework—significantly outperformed government-run CPs that initially adopted only the first component. COH-supported CPs achieved substantially higher percent positivity (32.5% vs. 9.5%) and viral load suppression rates (92.5% vs. 84.9%). After government sites integrated all three components, performance improved across all indicators. These results underscore the importance of full model fidelity in building trust, reducing HIV-related stigma, and delivering high-quality, person-centered care.

### CP’s responsiveness to COVID-19

The flexibility that enabled CPs to address gaps in HIV care also allowed them to adapt rapidly during the COVID-19 pandemic. CPs expanded their scope to provide COVID-19 prevention, contact tracing, and vaccination while continuing essential HIV and primary care services. HIV-related tools and strategies—such as index testing and stigma reduction messaging—were successfully adapted to the COVID-19 response. Faith leaders helped disseminate “Messages of Hope” to combat misinformation and vaccine hesitancy, contributing to high vaccine acceptance. These efforts demonstrate the model’s versatility and its capacity to support integrated health responses in times of crisis.

### Strengths and Limitations

This analysis has several strengths. It draws on four years of routinely collected program data, spanning pre- and intra-pandemic periods. The data are disaggregated by sex, enabling a focused understanding of men’s access to HIV services. Comparisons with government-supported CPs strengthen external validity by providing a benchmark for assessing the effectiveness of the intervention in a real-world setting. The CP model’s adaptability during the pandemic further underscores its operational resilience despite unprecedented challenges. However, there are limitations. First, the observational nature of the data limits causal inference. Second, PEPFAR testing indicators count tests conducted rather than individuals tested, which may lead to overestimation of unique clients tested and diagnosed. Third, the analysis lacks longitudinal tracking of individuals, which restricts insight into long-term retention and viral suppression. Finally, outcomes may be influenced by local contextual factors—such as leadership or community dynamics—limiting generalizability. To address this, replication efforts are underway in Zimbabwe, Kenya, South Sudan, and Côte d’Ivoire to assess the model’s effectiveness in diverse settings.

## Conclusion

The CoH CP model is a promising solution to improve engagement of men in HIV services. By leveraging trust, minimizing structural barriers, and embedding values-based care in service delivery, the model achieved substantial gains in HIV testing, treatment, and viral suppression among men. Based on these results, the Zambian government has scaled the model to over 105 CPs across seven provinces, and the World Health Organization has highlighted it as an evidence-based approach for engaging men in HIV care [31]. Future efforts should explore adaptation of this model to other public health challenges and evaluate its long-term cost-effectiveness compared to traditional facility-based care.

## Data Availability

All data produced in the present study are available upon reasonable request to the authors

## Acknowledgements

The authors would like to express their heartfelt gratitude to all Circle of Hope staff for their unwavering dedication and tireless efforts to provide high-quality HIV services to the people in their communities.

The authors would also like to thank Minesh Shah and Issac Zulu for their invaluable support and insightful feedback during the planning stages of this analysis. Your contributions have been instrumental in bringing this work to fruition. Finally, the authors would like to thank Mwayabao Jean Claude Kazadi and Olufunlola Adedeji from Catholic Relief Services who played a key role in developing and supporting the faith- and community-led Community Post model in its early stages.

## Disclaimer

The findings and conclusions in this report are those of the authors and do not necessarily represent the official position of the funding agencies.

## Data Availability Statement

The data that support the findings of this study are available from the corresponding author upon reasonable request.

